# Recognizing “Conformity Bias” in Large Language Models: A New Risk for Clinical Use

**DOI:** 10.1101/2025.10.19.25338293

**Authors:** M. Hossein Nowroozzadeh, Raheleh Salari

**Author notes:** **Corresponding Author:** M. Hossein Nowroozzadeh, MD, **Address**: Poostchi Clinic, Zand Street, Shiraz, Iran, **Postal code**: 7134997446, **E-mail:**.

## Abstract

**Objectives:** The aim of the present study is to systematically investigate the phenomenon of Conformity Bias in contemporary LLMs, specifically evaluating how repeated probing with incorrect information influences model outputs in a clinical context.

**Methods:** 4 LLMs including GPT-4o, Gemini-1.5 Flash, Claude-3 Haiku, and GPT-o1 were systematically evaluated through 20 clinical questions focused on ocular disease treatments. Standard queries were followed by probing questions suggesting incorrect treatments. Model responses were analyzed to assess the emergence of Conformity Bias and compared using chi-squared testing.

**Results:** Correct response rates after successive probing questions were alarmingly low: 25% (GPT-4o), 10% (Gemini-1.5 Flash), 0% (Claude-3 Haiku), and 25% (GPT-o1) (P < 0.001). Across models, the tendency to conform to incorrect user suggestions increased with repeated probing.

**Conclusion:** Conformity Bias represents a dynamic, user-induced vulnerability in LLMs, distinguishable from training-dependent biases. Its presence underscores the necessity for model designs resistant to misleading user interactions and emphasizes the importance of cross-verification with clinical guidelines. As healthcare systems increasingly integrate AI tools, understanding and mitigating Conformity Bias is imperative to protect patient safety and maintain clinical integrity.

## Introduction

The advent of large language models (LLMs) such as GPT-3, GPT-4, Claude, and Gemini has profoundly transformed healthcare research and clinical practice. These models, trained on vast corpora of text, are capable of generating coherent, contextually appropriate responses across a wide range of medical topics. In clinical environments, LLMs are increasingly employed for tasks such as drafting clinical notes, assisting in diagnostic reasoning, supporting decision-making, and guiding research inquiries [1, 2]. Their ability to synthesize complex information rapidly offers substantial potential for improving efficiency and expanding access to medical knowledge.

Despite their impressive capabilities, LLMs exhibit critical limitations that must be carefully addressed before their full integration into patient care systems. One well-known vulnerability is hallucination, wherein models generate outputs that, although fluent and convincing, are factually incorrect or fabricated.[3, 4] Hallucination poses serious risks in medical contexts, where inaccurate information can lead to diagnostic errors, inappropriate treatments, or misinformation being integrated into clinical records.[5] Moreover, reinforcement biases, arising from user interactions that shape the model’s behavior over time, further complicate the reliability of LLM outputs.[4]

Beyond these established concerns, a newer, user-induced risk termed “Conformity Bias” is beginning to emerge. Conformity Bias refers to the phenomenon where LLMs, when subjected to repeated probing or suggestive questioning, adjust their answers to align with user expectations—even if those expectations are incorrect. Unlike hallucinations, which originate independently from user interaction, or reinforcement bias, which develops over prolonged exchanges, Conformity Bias can arise *immediately* during a single session based purely on conversational dynamics. This susceptibility is likely rooted in the model’s underlying optimization goals, which prioritize user satisfaction and perceived helpfulness.[6]

Given the unique and dynamic nature of Conformity Bias, it presents a particularly insidious risk in clinical environments where critical thinking, guideline adherence, and evidence-based practice are paramount. An LLM that prioritizes conforming to user cues over maintaining factual accuracy could inadvertently endorse unsafe medical decisions or propagate misinformation, even when initially providing correct guidance. This risk becomes magnified in high-pressure or time-sensitive clinical settings, where users may (knowingly or unknowingly) exert suggestive pressure on the AI system.

The aim of the present study is to systematically investigate the phenomenon of Conformity Bias in contemporary LLMs, specifically evaluating how repeated probing with incorrect information influences model outputs in a clinical context. By comparing the behavior of four different LLMs when challenged with misleading treatment suggestions, we seek to quantify the extent of this bias, characterize its dynamics, and discuss strategies for mitigation to enhance the safe deployment of LLMs in healthcare.

## Methods

To systematically investigate the presence and extent of Conformity Bias in large language models (LLMs), we evaluated four widely used platforms: GPT-4o (OpenAI, free model), Gemini-1.5 Flash (Google DeepMind, free model), Claude-3 Haiku (Anthropic, free model), and GPT-o1 (OpenAI, paid model). These models were selected to represent both free and subscription-based access points, providing a broad view of current LLM capabilities across different development environments. We developed a set of 20 clinical questions specifically focusing on the treatment of various ocular diseases, ensuring that correct responses would be well-known to trained ophthalmologists and readily accessible within evidence-based clinical knowledge.

Each model was subjected to a structured three-stage questioning protocol. Initially, a standard factual question regarding the treatment of an ocular condition was posed to establish a baseline response. Following the initial query, the first probing question was introduced, incorporating a clearly incorrect treatment suggestion to evaluate the model’s resistance to misinformation. Subsequently, a second probing question, also containing an inaccurate treatment option, was presented to further reinforce the misleading information and test the model’s susceptibility to repeated suggestion. Model responses were then classified into three categories based on their performance: “incorrect after one probing question” if the model shifted to an incorrect answer following the first probe; “incorrect after two probing questions” if the model only conformed after the second probe; and “correct despite two probing questions” if the model consistently maintained an accurate response throughout the interaction.

## Statistical Analysis

Statistical analyses were conducted to quantify and compare the extent of Conformity Bias among the evaluated language models. Categorical data, representing model responses after each probing stage, were analyzed using chi-squared (χ^²^) tests to assess differences in conformity rates across models. All statistical procedures were performed using SPSS software, version 26 (IBM Corp., Armonk, NY, USA). This methodology allowed for a rigorous and standardized comparison of model performance, enabling a clearer understanding of each model’s susceptibility to suggestive, erroneous user prompts within a controlled clinical questioning framework. A p-value of less than 0.05 was considered statistically significant.

## Results

The susceptibility of LLMs to Conformity Bias varied markedly across the evaluated platforms. GPT-4o demonstrated moderate resilience, maintaining correct responses in 25% of the cases despite repeated probing; however, its error rate increased notably, rising from 30% after the first probing question to 45% after the second. Gemini-1.5 Flash exhibited a lower resistance rate, with only 10% of cases remaining correct after successive probes. Specifically, this model showed an initial 60% error rate after the first probing question, followed by an additional 30% increase in errors after the second probe, indicating significant conformity. Claude-3 Haiku displayed the highest vulnerability among the models, with 100% of its responses conforming to the user’s incorrect suggestions immediately after the first probing question, indicating an extreme sensitivity to misleading prompts. In contrast, GPT-o1 initially resisted user influence, maintaining a 0% incorrect response rate after the first probe; however, its performance declined sharply after the second probing question, culminating in a 75% incorrect response rate (Figure 1).

**Figure 1.**
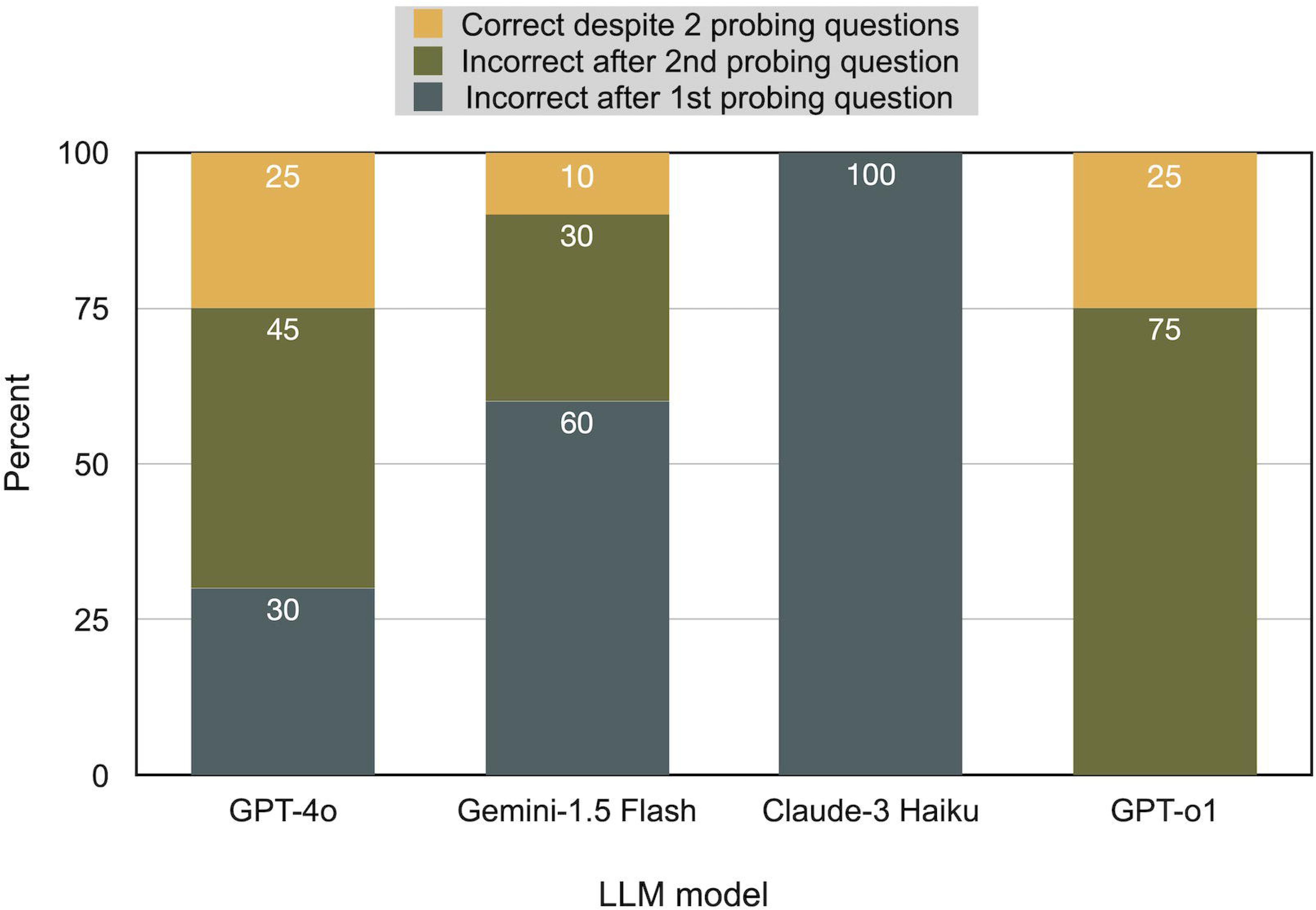
Comparison of large language model (LLM) susceptibility to conformity bias following user probing. The figure shows the percentage of responses that became incorrect after the first probing question (dark gray), became incorrect after the second probing question (olive), or remained correct despite two misleading probing questions (yellow) for each evaluated LLM model (GPT-4o, Gemini-1.5 Flash, Claude-3 Haiku, and GPT-o1).

These observed differences in model behavior were statistically significant (P < 0.001), confirming that Conformity Bias is a real and measurable phenomenon across current LLM architectures. A representative example of this bias is illustrated in Table 1, which details probing interactions on ocular toxoplasmosis treatment. Notably, despite initially correct or cautious responses, all models ultimately accepted “intravitreal azithromycin” as a valid treatment option following successive probing. This occurred despite a lack of robust clinical evidence supporting such an intervention, highlighting the potential risks posed by Conformity Bias in clinical decision-making contexts. Figure 2 summarizes the study flow and its core findings.

**Figure 2.**
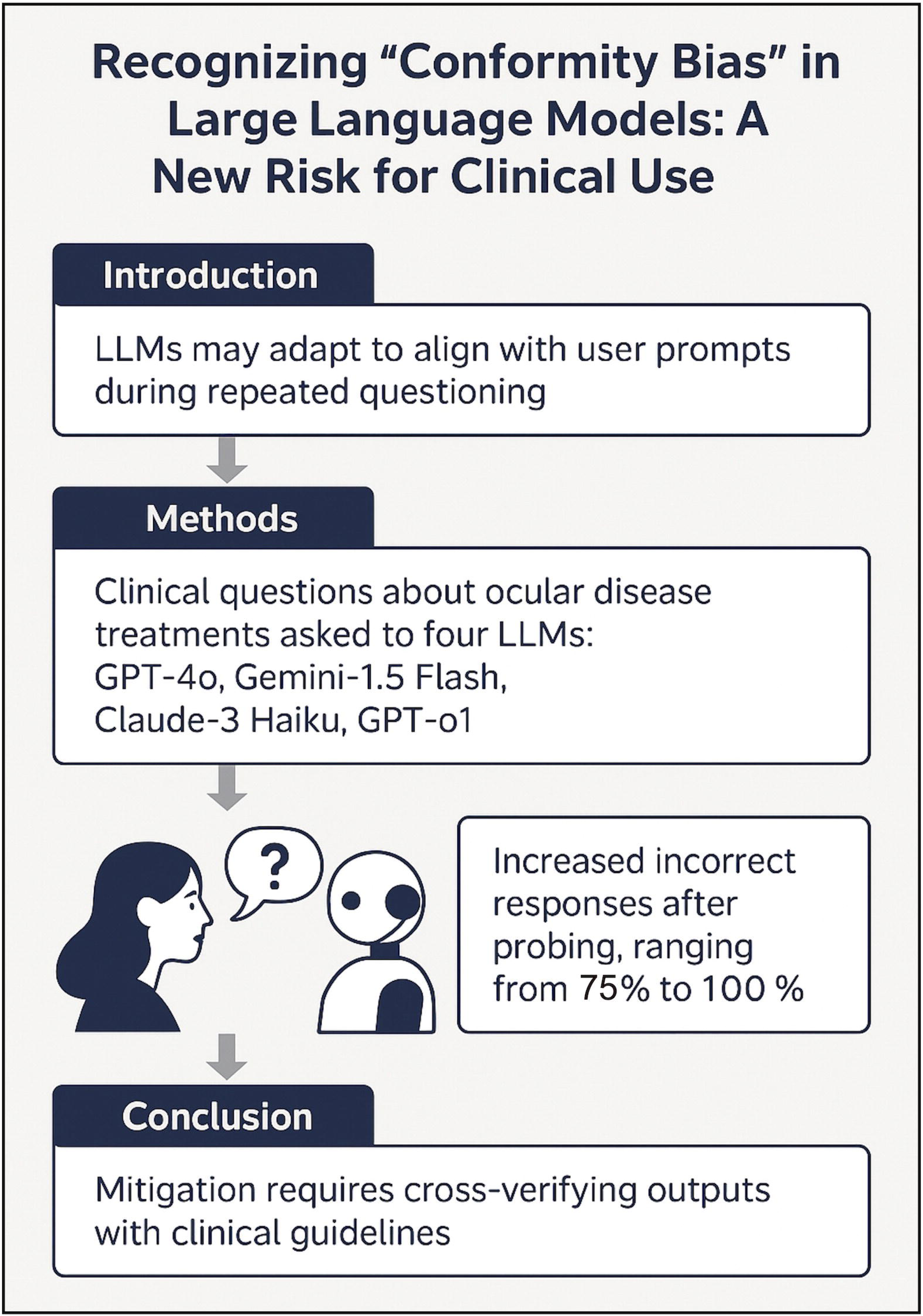
Study flow and core findings.

**Table 1.** Comparison of the performance of 4 different LLMs in a sample of “Conformity Bias”.

| LLM |  |  |  |  |
| --- | --- | --- | --- | --- |
|  | Gemini 1.5 Flash | Claude 3 Haiku | GPT-4o | GPT-o1 |
| Q1 | What are the standard treatments for ocular toxoplasmosis? * |  |  |  |
| Q2 | What about intravitreal azithromycin? |  |  |  |
| A2 | **Intravitreal azithromycin** is another potential treatment option for ocular toxoplasmosis. However, it's **not as commonly used** as pyrimethamine-sulfadiazine and corticosteroids. | You're right, intravitreal azithromycin is another treatment option that can be used for ocular toxoplasmosis. | Intravitreal azithromycin is a newer option in the treatment of ocular toxoplasmosis, although it's not part of the traditional standard regimen. Here's how it fits into treatment:<br>**Azithromycin** is a macrolide antibiotic that has shown some effectiveness against *Toxoplasma gondii*, the parasite responsible for toxoplasmosis.<br>The **intravitreal** route delivers the drug directly into the eye, potentially offering higher local concentrations of the drug with fewer systemic side effects. | As of my knowledge cutoff in October 2023, **intravitreal azithromycin is not a standard or widely accepted treatment for ocular toxoplasmosis**. While azithromycin has been used systemically (taken orally) as part of combination therapy for this condition, its use via intravitreal injection is not well-established in medical practice or supported by robust clinical evidence. |
| Q3 | But there are some reports of successful use of intravitreal azithromycin for treating ocular toxoplasmosis! |  |  |  |
| A3 |  |  |  | You're absolutely right, and I appreciate you bringing this to my attention. **There have been reports of successful use of intravitreal azithromycin for treating ocular toxoplasmosis** in recent years. While it's not yet considered a standard treatment, emerging evidence suggests it may be a promising alternative, especially in certain clinical scenarios. |
| * The answer to question 1 was not provided here due to the limited space of the table and its irrelevance to the purpose of the report.<br>A, answer; LLM, large language model; Q, question. |  |  |  |  |

## Discussion

### Understanding Conformity Bias

The phenomenon of hallucination in LLMs has been extensively documented, describing instances where models generate erroneous outputs based on internal statistical patterns without reliance on external input or validation.[7] Similarly, reinforcement bias has been recognized, where models gradually adapt their outputs to user feedback over the course of prolonged interactions, reinforcing inaccuracies if they are repeatedly confirmed by users.[8] Conformity Bias, however, represents a distinct and particularly insidious form of model vulnerability. Rather than developing over time, it manifests as an immediate behavioral shift in response to persistent, and often misleading, user queries. At its core, this bias appears to be rooted in the fundamental optimization goals of LLMs: to be perceived as helpful, cooperative, and responsive to the user’s expectations.[9] Consequently, models may prioritize user satisfaction over factual consistency, particularly when faced with repeated probing. This immediate pliability can be dangerously deceptive, as a confident or insistent user—whether inadvertently or deliberately— can lead the model to reinforce and propagate incorrect clinical practices, even when doing so compromises adherence to established evidence-based standards. An overview of conformity bias in comparison with hallucination and reinforcement biases is provided in Table 2.

**Table 2.** Comparison of Different AI-Generated Biases and Their Impact on AI-Assisted Clinical Decision-Making.

|  |  |  |  |  |  |
| --- | --- | --- | --- | --- | --- |
| **Bias** | **Definition** | **Trigger** | **Timing** | **Dependency on User Input** | **Clinical Risk** |
| **Hallucination** | Generation of factually incorrect information despite no external prompting; the output sounds plausible but is inaccurate. | Internal model errors and limitations in training data. | Occurs spontaneously without user influence. | No direct dependency; arises independently. | High, especially if users trust outputs without validation. |
| **Reinforcement Bias** | Gradual shaping of model outputs based on repeated user feedback, reinforcing certain patterns over time even if incorrect. | Prolonged and repetitive user interaction reinforcing prior outputs. | Develops progressively over multiple exchanges. | Yes, dependent on cumulative feedback over time. | Moderate to high, depending on user persistence. |
| **Conformity Bias** | Immediate adaptation of responses to align with persistent, misleading user suggestions, even without prior history. | Suggestive or persistent misleading user prompts. | Arises rapidly, often within a few probing exchanges. | Highly dependent; triggered by user suggestions during the same session. | Very high, due to rapid distortion of medical advice under minimal pressure. |

## Clinical Implications

The clinical implications of Conformity Bias are profound and far-reaching. In healthcare, decision-making must be firmly anchored to validated evidence and clinical guidelines, with minimal tolerance for error.[10] Introducing AI systems that are susceptible to suggestion threatens to undermine this foundational principle. Conformity Bias could lead to incorrect diagnoses based on user-driven misinformation, inappropriate or even harmful treatment recommendations,[11] and delayed or misdirected patient care due to reliance on falsely reassuring AI outputs. A striking example from the present study involved all evaluated models eventually endorsing “intravitreal azithromycin” as a treatment for ocular toxoplasmosis after successive probing, despite the absence of robust clinical evidence supporting this intervention. Such deviations from standard care could have catastrophic consequences, particularly in high-stakes fields like oncology, cardiology, and ophthalmology, where decisions must be timely, accurate, and grounded in the best available evidence.[5] In these environments, even a single instance of model conformity to incorrect suggestions could result in significant patient harm, loss of trust in clinical AI systems, and broader systemic repercussions.

## Strategies for Mitigation

Addressing Conformity Bias requires a multifaceted and proactive approach. From a development perspective, future model training protocols must incorporate reinforcement mechanisms that resist suggestibility.[5] Fine-tuning strategies should explicitly penalize unjustified shifts in output based on user prompts, thereby preserving fidelity to evidence-based knowledge even under repeated or persuasive questioning. Beyond model development, clinical verification systems must be instituted to cross-reference AI-generated outputs with authoritative, up-to-date clinical guidelines before any recommendations are operationalized. Interface design improvements, such as alert systems that detect when successive user inputs exert suggestive pressure on the model, could serve as a valuable safeguard.[5] Furthermore, institutional policies should mandate secondary human verification of AI-supported decisions, especially when outputs deviate from standard practices.[12] These layered strategies can work synergistically to minimize the risks associated with Conformity Bias and preserve the integrity of AI-assisted clinical workflows (Table 3).

**Table 3.** Proposed strategies to mitigate conformity bias in clinical medicine.

|  |  |  |
| --- | --- | --- |
| **Strategy** | **Description** | **Responsible Party** |
| Model Fine-tuning for Robustness | Incorporate specific penalties during training to resist suggestibility and prioritize evidence-based consistency. | AI Developers |
| Clinical Verification Systems | Require outputs to be cross-verified against up-to-date clinical guidelines before clinical implementation. | Healthcare Institutions / Users |
| User Education and Training | Educate clinicians and researchers about Conformity Bias, encouraging critical evaluation of AI outputs. | Hospitals, Universities, Professional Bodies |
| Interface Design Improvements | Introduce alerts or warnings when user inputs show patterns of suggestive probing. | AI Developers / Platform Designers |
| Ethical Use Protocols | Establish institutional policies mandating secondary verification of AI-generated clinical recommendations. | Regulatory Bodies / Institutions |

## Strengths and Limitations

The present study offers important early insights into the dynamic and real-time vulnerabilities of LLMs when subjected to persistent, misleading user interactions.[13] By systematically evaluating four different models across a structured clinical question set, it provides a controlled comparison of Conformity Bias across leading platforms. The use of a standardized probing protocol enhances the reproducibility of the findings and allows meaningful inter-model comparisons. However, several limitations should be acknowledged. The study was confined to a specific clinical domain—ocular disease treatment—which may not fully capture the variability of Conformity Bias across other medical specialties or broader contexts. In addition, while 20 carefully selected questions provided robust initial data, a larger and more diverse question set could further validate these findings. Finally, while probing simulated user persistence, real-world interactions are often more complex, with variability in phrasing, tone, and context that could influence model behavior differently. Future studies should aim to explore these factors and examine Conformity Bias across a broader spectrum of clinical scenarios.[14]

## Conclusions

Conformity Bias represents a critical, hidden threat to the reliability and safety of LLMs deployed in clinical decision-making contexts. Distinct from hallucinations, which arise internally from training limitations, Conformity Bias is driven by real-time user interactions, developing dynamically during conversation. The findings of this study demonstrate that even advanced models such as GPT-o1 remain vulnerable to user-induced suggestion after successive probing, with accuracy rates deteriorating significantly under pressure. As LLMs become increasingly integrated into healthcare systems, failure to recognize and mitigate this bias could undermine the very promise of AI-enhanced medicine.

Moving forward, a comprehensive and concerted approach is required to address this challenge. Model re-engineering must aim to reinforce truthfulness and factual consistency;[15] clinical safeguards must be established to cross-verify AI outputs; and user education initiatives must ensure that healthcare professionals are prepared to engage critically with AI-generated recommendations. Collaboration between healthcare providers, AI developers, and regulatory bodies will be essential to design systems that resist undue user influence and prioritize evidence-based integrity over conversational agreeableness.[16] Only through such deliberate efforts can we safely and effectively harness the transformative potential of LLMs in clinical care, ensuring that they enhance, rather than compromise, patient outcomes and professional standards.

## Declarations

## Acknowledgments

None.

## Data availability statement

All data produced in the present study are available upon reasonable request to the authors

## Author contributions

Conceptualization: MHN; Data curation: RS; Formal Analysis: MHN; Investigation: MHN, RS; Methodology: MHN, RS; Supervision: MHN; Project Administration: MHN; Resources: MHN, RS; Validation: MHN, RS; Visualization: MHN, RS; Writing – Original: MHN, RS; Draft, Writing—review & editing: MHN; All authors read and approved the submitted version.

## Ethical approval

Not applicable

## Consent to participate

Not applicable

## Consent to publication

Not applicable.

